# Allogeneic Hematopoietic Cell Transplants as Dynamical Systems: Effect of Early-Term Immune Suppression Intensity on Long-Term T Cell Recovery

**DOI:** 10.1101/2022.04.22.22274154

**Authors:** Viktoriya Zelikson, Roy Sabo, Myrna Serrano, Younus Aqeel, Savannah Ward, Taha Al Juhaishi, May Aziz, Elizabeth Krieger, Gary Simmons, Catherine Roberts, Jason Reed, Gregory Buck, Amir Toor

**Affiliations:** Department of Internal Medicine, Virginia Commonwealth University, Richmond, VA; Department of Biostatistics, Virginia Commonwealth University, Richmond, VA; Department of Microbiology and Immunology, Virginia Commonwealth University, Richmond, VA; Department of Pharmacy, Virginia Commonwealth University, Richmond, VA; Department of Pediatrics, Virginia Commonwealth University, Richmond, VA; Department of Physics, Virginia Commonwealth University, Richmond, VA

**Keywords:** Dynamical systems, Graft vs. host disease, hematopoietic cell transplantation, T cell repertoire, Complex numbers, immunoablative conditioning

## Abstract

Reduced intensity conditioning (RIC) is fraught with risk for disease relapse. This may be overcome by donor T cell alloreactivity. Reducing the duration of intense immune suppression in the early term following transplantation may create an immunologic environment favoring rapid T cell reconstitution to influence longer term transplant outcomes. Twenty-six patients were adaptively randomized based on donor-derived T cell recovery, between 2 different dosing schedules of mycophenolate mofetil (MMF): MMF for 30 days post-transplant, with filgrastim for cytokine support (MMF30 arm, 11 patients), or MMF for 15 days post-transplant, with sargramostim (MMF15 arm, 15 patients). All patients were treated with anti-thymocyte globulin at a dose of 1.7 mg/kg/day from day - 9 through day -7 and total body irradiation, 450 cGy given in 3 fractions. Patients were well matched between the study arms and underwent HLA matched related and unrelated donor hematopoietic cell transplantation (HCT). The MMF15 arm demonstrated superior T cell recovery in the first month. This difference persisted through the first year for total T cells, T cell subsets and NK cells. T cell repertoire tended to be more diverse in the MMF15 arm. The long term superior immune recovery in the MMF15 arm is consistent with a disproportionate impact of early interventions in HCT. Clinically, shorter course MMF post-transplant was not associated with increased risk of acute or chronic graft vs. host disease (GVHD), and relapse and there was a trend toward improved overall survival in the MMF15 arm. Reducing the duration of intense immunosuppression in the early term and the use of sargramostim following allogeneic HCT is feasible and leads to superior long-term T cell recovery. This regimen should be studied to improve immune recovery in large cohorts of patients undergoing HCT with RIC.

## Introduction

Allogeneic hematopoietic cell transplantation (HCT) is conventionally modelled as a stochastic system, where recipient, donor and disease characteristics determine clinical outcomes.^1^ The probabilities of optimal outcomes, such as regimen related toxicity, graft versus leukemia (GVL) effect, graft versus host disease (GVHD) and infections are governed by the conditioning intensity, GVHD prophylaxis, disease control and human leukocyte antigen (HLA) matching of donors and recipients. Conventional clinical trial design lends itself to studying the consequences of defined interventions in a limited number of these parameters – for example, comparing myeloablative vs. reduced intensity conditioning (RIC) for transplanting patients with acute myelogenous leukemia (AML),^2^ or the use of bone marrow vs. peripheral blood stem cells (PBSC) for unrelated donor transplantation.^3^ This has been a remarkably successful paradigm which has over the years led to the evolution of the field from a perilous, seldom used option, to a routinely applied therapeutic modality. Nevertheless, at an individual patient level, outcomes remain stochastic with little predictability of complications of therapy in transplant recipients. Close clinical follow up and laboratory monitoring for variables such as measurable residual disease and chimerism still provide the best tools for early intervention and achieving optimal outcomes.^4^ However these tend to require a considerable lead time to be interpretable and are generally not informative in the early term post HCT.

Major causes of treatment failure after an allograft are relapsed malignancy and non-relapse mortality (NRM), with a reciprocal relationship between conditioning intensity and GVHD prophylaxis for both outcomes. RIC reduces NRM, but relapse risk is higher in some malignancies less susceptible to GVL effects, while intense GVHD prophylaxis may increase infection and relapse risk, and inadequate immune suppression increases GVHD likelihood through the different strata of histocompatibility between donors and recipients. The current state of science is to apply immunosuppression in equal measure across donors with a given level of HLA compatibility.^5^ As a consequence considerable variation is observed in immune recovery post-transplant, and subsequently in clinical outcomes.^6^ Complex trial design addressing multiple aspect of transplant immunobiology are therefore needed to increase the efficiency with which clinical trials lead to therapy optimization.

Previous work has demonstrated that immune reconstitution following HCT has many characteristics of a dynamical system; systems which may be modelled with mathematical precision.^7 8^ As an example, T cell recovery post-transplant may be modelled utilizing equations of growth, where cell proliferation occurs as a logistic function of time. Furthermore, in an antigen affinity driven logistic expansion model of individual T cell clonal growth, Power law distribution of the T cell clonal frequencies is observed.^9^ This is similar to the clinical studies of T cell receptor beta repertoire in normal donors and recipients following transplantation, where an expanding repertoire is documented in the months following transplantation.^10 11^ Further evidence of dynamical behavior comes from the observation that early interventions have a lasting impact on transplant outcomes, for example the use of post-transplant cyclophosphamide which mitigates chronic GVHD.^12 13 14^ Chronic GVHD risk is also mitigated using anti-thymocyte globulin (ATG), particularly following HLA matched unrelated donor HCT.^15 16^ However, ATG administration delays T cell recovery, especially CD4+ helper T cell reconstitution^17 18 19^ and may impact relapse and viral infection risk. The timing of administration of ATG for in vivo T cell depletion alters the dynamics of T cell recovery post-transplant and allows superior T cell recovery if given early in the conditioning regimen.^8 20^ Further, given the dynamical systems nature of immune recovery, the T cell reconstitution may be further enhanced by adjustment in post-transplant immune suppression.

Post-transplant GVHD prophylaxis in conventional regimens includes a calcineurin inhibitor (CNI) given with a cell cycle active agent such as methotrexate or mycophenolate mofetil (MMF).^21^ MMF is generally given over a month, marking the period of intense post-transplant immune suppression. The duration of MMF administration comes from clinical trials following the original canine experiments where CNI+MMF was the optimal immune suppression for non-myeloablative (NMA) conditioning with 2-Gray (Gy) total body irradiation (TBI) and HCT.^22^ This concept combined with related work demonstrating engraftment across major histocompatibility antigen barriers with anti-T cell antibodies^23^ formed the basis of immunoablative, RIC regimen combining 4.5-Gy TBI with early administration of ATG. ^20 24^ The adaptively randomized trial reported here, tested the effect of administering intense post-transplant immune suppression for different duration based on donor-derived T cell count at 8 weeks post-transplant, in patients conditioned with the ATG and 4.5-Gy TBI regimen (ATG-TBI).^25^ T cell recovery results from the two study arms are reported, as are the clinical outcomes.

## Methods

### Patients and adaptive randomization scheme

The reported study was an adaptively randomized, phase 2, open-label, trial (NCT02593123), approved by the Virginia Commonwealth University IRB (MCC-14-10739). Inclusion criteria included high risk or recurrent hematologic malignancy, and availability of 8/8 or 7/8, high resolution HLA-A, -B, -C, and DRB1 matched, related (MRD) or unrelated donor (MUD). Allocations to the study and control arms was stratified based on, 1: lymphoid versus myeloid malignancy, and 2: donor type MRD vs. MUD (Table 1). An adaptive randomization scheme was utilized to increase the probability of optimal patient outcomes,^26^ with the stratification algorithm being a doubly adaptive biased coin design (DBCD) coupled with optimal allocation of continuous outcome,^27^ in this case donor-derived CD3+ (ddCD3) T cell count at 8 weeks post-transplant. This was calculated by the following formula: *ddCD3=absolute CD3+ cell count * fraction donor chimerism in T cells* (Equation 1). This surrogate marker was chosen because a higher ddCD3 count on day-60 was previously shown to be associated with improved outcomes in an earlier cohort of the ATG-TBI conditioned patients with GVHD prophylaxis as delivered in the control arm. ^25^ Initial enrollment had an equal number of patients per arm, with treatment allocation recalculated on a rolling basis.

**Table 1.** Patient demographics.

| Demographics | MMF 15 | MMF 30 | P (FET) |
| --- | --- | --- | --- |
| N | 15 | 11 |  |
| Age (median) | 61.5 (50-68) | 56.7 (42-69) | 0.25 (Wilcoxon) |
| Gender (F) | 6 (40%) | 6 (54.5%) | 0.69 |
| KPS (90-100) | 5 (33.3%) | 3 (27.2%) | 1 |
| Disease Type (Myeloid) | 6 (40%) | 3 (27.2%) | 0.68 |
| AML | 5 (30%) | 2 (18.2%) | 0.66 |
| CLL | 4 (26.6%) | 1 (9.1%) | 0.36 |
| CML | 0 | 1 (9.1%) | 0.42 |
| MDS | 1 (6.7%) | 1(9.1%) | 1 |
| MM | 2 (13.3%) | 2 (18.2%) | 1 |
| NHL | 3 (20%) | 4 (36.4%) | 0.41 |
| HLA Match (8/8,10/10)* | 14 (93.3%) | 8 (72.7%) | 0.27 |
| HLA-MRD | 6 (40%)** | 2 (18.2%) | 0.39 |
| Disease Risk Index (DRI) |  |  |  |
| Low | 3 | 3 | 1 |
| Intermediate | 8 | 6 | 1 |
| High | 4 | 2 | 0.67 |
| Gender Mismatch |  |  |  |
| M (R) x F (D) | 1 | 1 | 1 |
| M (R) x M (D) | 8 | 4 | 0.45 |
| F (R) x M (D) | 3 | 2 | 1 |
| F (R) x F (D) | 3 | 4 | 0.40 |
| CMV Seropositivity |  |  |  |
| + (R) x + (D) | 6 | 4 | 1 |
| + (R) x - (D) | 3 | 1 | 0.61 |
| - (R) x + (D) | 0 | 3 | 0.06 |
| - (R) x - (D) | 6 | 3 | 0.68 |
| Blood Type (mismatched) | 6 | 3 | 0.68 |
| Engraftment time (median) | 14 | 11 | **0.02** (Wilcoxon) |
| GCSF- mobilized PBSC | 15(100%) | 10(91%) | 0.423 |
| CD34 Dose (10<sup>6</sup>)(mean/SD) | 4.52 ± 1.71 | 4.81±2.29 | 0.86 (Wilcoxon) |

\*MMF15 had 1 MMUD transplant mismatched at the HLA-DQ locus. MMF30 had 3 MMUD transplant mismatched with 1 patient mismatched at the A + DQ loci, and 2 patients mismatched at the A loci. \*\* One patient with a dizygotic twin donor (F to M).

### Conditioning and study schema

All patients were conditioned with rabbit ATG (Thymoglobulin, Sanofi-Aventis, Bridgewater, NJ) at a dose of 1.7 mg/kg/day from day -9 through day -7 (total dose 5.1 mg/kg), and 1.5 Gy TBI administered twice daily on day -1 and once on day 0 for a total dose of 450 cGy given in 3 fractions. ^24 25^ Patients randomized to the investigational cohort (MMF15) received mycophenolate mofetil (MMF) dosed at 15 mg/kg every 12 hours from day 0 to day 15 and administered subcutaneous granulocyte-macrophage colony stimulating factor (GM-CSF) (Sargramostim, Partner Therapeutics, Lexington, MA) 250 mcg/m^2^/day from day 4 until hematopoietic reconstitution; those on the control arm (MMF30) received MMF dosed at 15 mg/kg every 12 hours from day 0 to day 30 and received subcutaneous granulocyte colony stimulating factor (G-CSF) (Filgrastim, Amgen, Thousand Oaks, CA) 5mcg/kg/day from day 4 through hematopoietic reconstitution. GVHD prophylaxis consisted of tacrolimus from day - 2 through day 90 followed by a two to three month long tapering schedule. In the first month the tacrolimus levels were maintained in a 10-15 ng/mL range dropping down to 8-12 ng/mL in the next two months generally. Patients in the investigational cohort were also given inhaled fluticasone twice daily from day 4 until discontinuation of GM-CSF to minimize pneumonitis risk.

### Engraftment and Immune Reconstitution

Circulating absolute monocyte, lymphocyte (ALC), and neutrophil count (ANC) were determined as a part of routine complete blood counts using a hematology analyzer (XN-9000; Sysmex, Lincolnshire, IL). CD3+, CD3+/CD4+, CD3+/CD8+, CD19+, and CD56+/16+ cells were measured on days 30, 60, 90, 180, 365, and 500 using a FACSCanto II flow cytometer (BD Biosciences, San Jose, CA). Donor T cell chimerism were also measured at these time points using PCR for short tandem repeats on DNA isolated from T cells isolated using anti-CD3 immunomagnetic beads. Donor-derived T cell and T cell subset counts (ddCD3, ddCD4, and ddCD8) were determined as noted above in Equation 1.

### T cell receptor beta sequencing

T cell receptor beta sequencing was performed using genomic DNA from transplant recipients obtained on approximately days 30 and 100 using the Adaptive Biotechnologies® immunoSEQ® human T-cell receptor beta (hsTCRB) Kit (Adaptive Biotechnologies, Seattle, WA). Genomic DNA was isolated from cryopreserved cell pellets obtained by processing 3 mL of blood, using DNeasy Blood & Tissue Kit and concentrated using Zymo Research DNeasy Blood & Tissue Kit following manufacturer’s instructions. DNA concentration was determined using Promega QuantiFluor dsDNA System kit and DNA integrity was determined by electrophoresis using 0.8% agarose gel. Four genomic DNA replicates per sample, each containing on average approximately 4.0 μg were independently amplified using Qiagen 2× Multiplex PCR Master mix with proprietary immunoSEQ® hsTCRB primers. The amplicons were diluted (1:4) and submitted to a second round of PCR amplification to add sample-specific barcodes and Illumina adapters to each PCR replicate. The final amplicons were combined into 2 pools (containing 23 and 17 DNA samples respectively) and cleaned up with magnetic beads provided in the kit. Pools were run on the Sage Science BluePippin DNA size selection system to remove primer dimers. A BluePippin 2% dye-free agarose gel cassette was used to capture DNA in a target range of 240-400 base pairs. After completion of size selection ∼50 µl of size selected libraries were removed from the elution wells, cleaned, and concentrated using a 1.8X Agencourt AMPure XP magnetic beads. Cleaned-size selected DNA libraries were eluted in 28 µl of Takara DNA suspension buffer. Size selection was assessed using Agilent Bioanalyzer High Sensitivity DNA Chip. Final DNA libraries were quantified using KAPA Library Quantification Kit. Sample pools were denatured with 0.2 N NaOH, diluted using Illumina Hybridization Buffer to 10pM and combined with 5% PhiX control, following Illumina guidelines. Samples were sequenced on the Illumina MiSeq instrument using MiSeq Reagent Kit v3 (150-cycle). The raw sequencing data were uploaded to Adaptive Biotechnologies immunoSEQ® ANALYZER and processed using the company’s proprietary pipeline. Results for each DNA sample were reported after merging data from all four replicates. Unique T cell receptor beta (TRB) gene rearrangements and templates in these samples were quantified and Simpson’s clonality determined (ImmunoSeq Analyzer 3.0).^28^

### T cell repertoire vector analysis

Conventional immune reconstitution studies look at T cell recovery and TRB sequencing information as distinct entities; however, these are two aspects of the same entity, the T cell repertoire, which evolves over time following HCT. The CD3+ cell count measures the number of circulating T cells, while the TRB sequencing measures the clonal makeup of the circulating T cells, each identified by its unique T cell receptor (TCR) variable, diversity and joining segment recombined sequence. Furthermore, each T cell clone may be modelled as a vector quantity because it has *magnitude* (clonal frequency) and *direction* (antigen specificity). ^9 29^ The unique T cell clones taken together constitute a multicomponent T cell repertoire vector (**Supplementary Figure 1A**). Clinical outcomes may be proportional to the overall impact of this vector. So, to understand the combined effect of T cell recovery on the clinical outcomes following transplantation, circulating CD3+ cell count and T cell clonality measures (such as the number of TCR rearrangements) may be modelled together as a *T cell repertoire vector*. Vector quantities such as the aforementioned T cell recovery measures, can be modelled in the complex number plane. Real numbers (*x*) in ordinary usage to measure quantities, occur on a number line (…-3, -2, -1, 0, 1, 2, 3,…). Complex numbers (*z = x + iy*), on the other hand, are mapped to a 2-dimensional plane, where the real component (*x*) is on the x-axis and the imaginary component 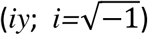 is on the Y-axis (**Supplementary Figure 1B**). Complex numbers are used to depict vectors using a single complex-valued number, *z*, instead of matrices with different aspect measures. The T cell repertoire may be similarly considered, where the circulating CD3+ T cell count at any given time *t*, is plotted against a TRB clonality measure, such as productive VDJ rearrangements at that time. The former measure gives an estimate of the sum of all T cell clonal frequencies, the *magnitude* of the T cell repertoire vector at time *t*, while the latter gives the number of T cell clones, the potential range of antigen specificities and thus, confers a *quality* or *direction* to the T cell number. This vector representation simultaneously accounts for both the quantitative (T cell count) and qualitative (TRB clonality) aspects of the T cell repertoire at any given point in time. T cell repertoire vectors were plotted at days 30 and 100 for each patient utilizing the absolute CD3+ cell count (μL^-1^) *1000 to get CD3+ cell counts mL^-1^. Adjusted for 3 mL this gave the potential number of T cells in the sample that the genomic DNA was extracted from. These T cell counts were plotted against the number of productive TRB gene rearrangements present at each time point to give complex valued T cell repertoire vector values.

### Statistical Analysis

Cell count differences between the two study arms were assessed with the Wilcoxon rank sum test. Clinical outcomes were analyzed using Cox proportional hazards with a univariate model only including the treatment arm (MMF15 vs MMF30). Multivariate models with the added variables of donor type (MRD vs MUD) and recipient age were applied to OS, NRM, and all forms of GVHD. Progression-free survival (PFS), relapse, and donor lymphocyte infusion (DLI)-free relapse-free survival outcome analyses were adjusted for treatment arm, donor type, and disease (myeloid vs lymphoid malignancy). In all Cox proportional hazards analyses involving GVHD, patients were censored at the date of relapse (competing risk) or their first DLI. Cumulative incidence of relapse was similarly censored at the time of NRM and DLI. Cumulative incidence analysis was used to compare both moderate-severe and severe cGVHD, with relapse as a competing event. Gray’s test was used to determine significance of differences observed between the two arms. GVHD was graded based on the National Institutes of Health consensus criteria.

The impact of various immune cell populations on clinical outcomes was analyzed with Cox proportional hazards analyses. Due to the small study size, all patients were pooled for this analysis and the study arm was used in the multivariate analysis to account for this variable’s effects. Cell counts were considered as continuous variables. Multivariate models including immune cell populations, included trial arm and donor status (MRD vs MUD), with the exception of relapse, and DLI-free, relapse-free survival which included trial arm and myeloid vs lymphoid disease distinction to parallel the other Cox analyses in this study.

## Results

### Clinical Outcomes

The two study arms were well matched for various study variables (Table 1). The median engraftment time was shorter in the MMF30 arm, which received GSCF (MMF15, day 14 vs. MMF30, day 11, p=0.0186). Patients in the MMF30 (GCSF) arm achieved a higher ANC (p=0.0384) and a faster maximum growth rate for monocytes (p=0.0714), (**Supplementary Table 1**). All patients in the study cohort had myeloid engraftment, median granulocyte chimerism was 100% donor-derived at days 30, 60 and 90, with 1 instance of graft failure in the control arm (FET; p=0.423). There was a trend toward improved OS in the MMF15 arm, though DLI-free, relapse-free survival, relapse and NRM were not different (**Figure 1**). Of the MMF15 patients who relapsed, 60% survived, compared to none in the MMF30 arm. DLI were used for, mixed chimerism (MMF15, 1 vs. MMF30, 0), relapse (4 v. 2), persistent disease (0 vs. 1), or graft failure (0 vs. 1), with no difference in overall DLI use between the two arms (MMF15, 5 vs. MMF30, 4, p=1). There was a trend toward increased aGVHD grade 2-4 in the MMF30 arm and conversely a trend toward increased moderate-severe cGVHD in the MMF15 arm (**Supplementary Figure 2**). There was no difference in cumulative incidence of relapse and cGVHD between the two arms (**Supplementary Figure 3**).

**Figure 1.**
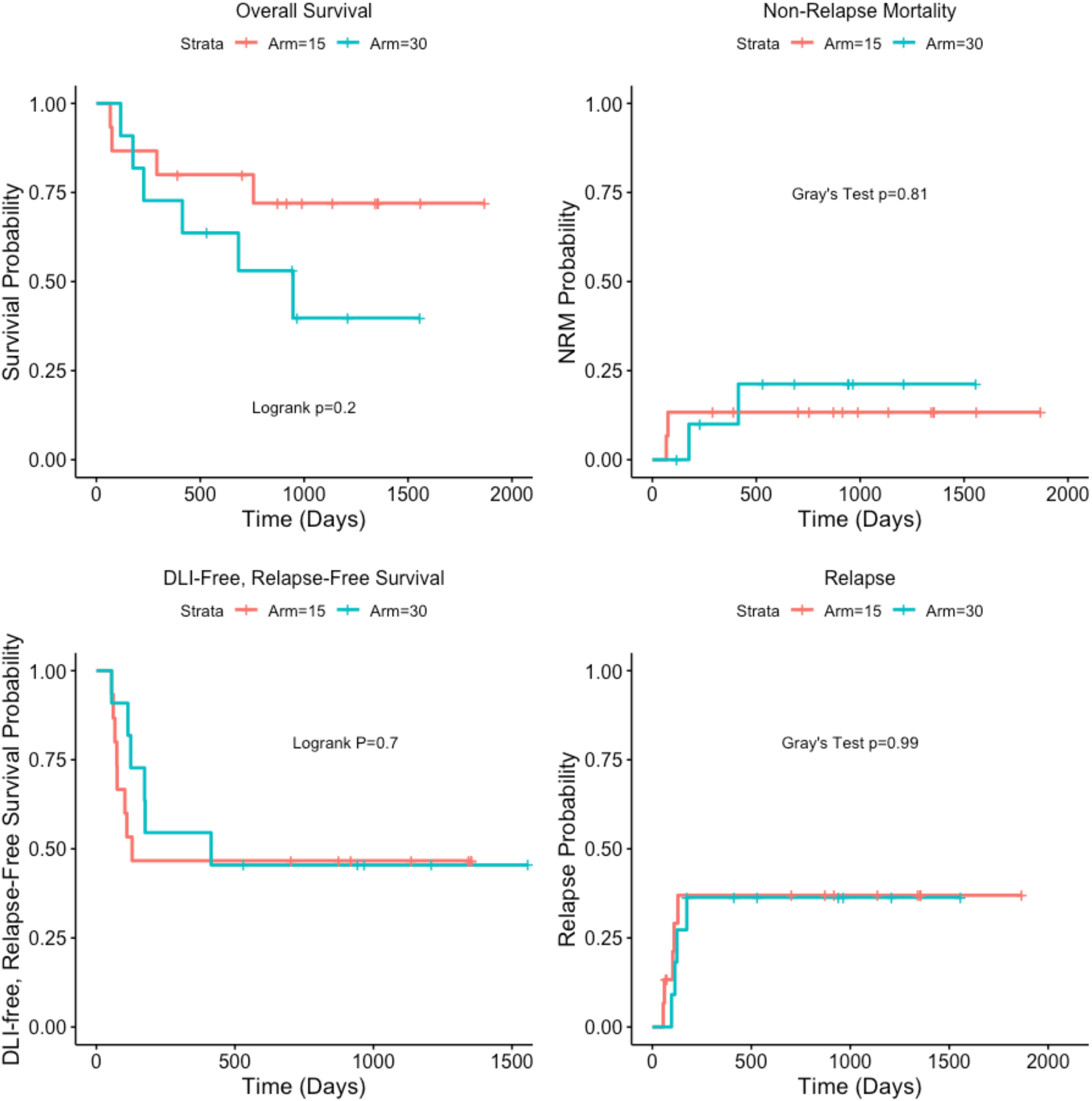
Clinical outcomes following HCT. Relapse, and NRM are competing events. Log Rank and Grays test were used for calculating the significance of difference between study arms.

### Early- and Long-term Immune Reconstitution

There was overall superior T cell recovery in the MMF15 arm; T cell recovery on day-30 was significantly higher in the MMF15 arm (**Figure 2**). Further, this impact was carried through beyond day 180 with the average CD3+ cell count being approximately two times higher in the MMF15 cohort, regardless of donor type. Though the randomization involved ddCD3 cell count on day 60, with a greater number of patients randomized to the study arm, there was no significant difference at day 60 in any T cell subset studied, underscoring that superior T cell recovery is not an artefact of the adaptive randomization process. CD3+4+ and CD3+8+ T cells were also consistently higher in the MMF15 arm, remaining 2-3 times higher for later time points regardless of the donor type. NK cells also showed superior reconstitution in the MMF15 arm.

**Figure 2.**
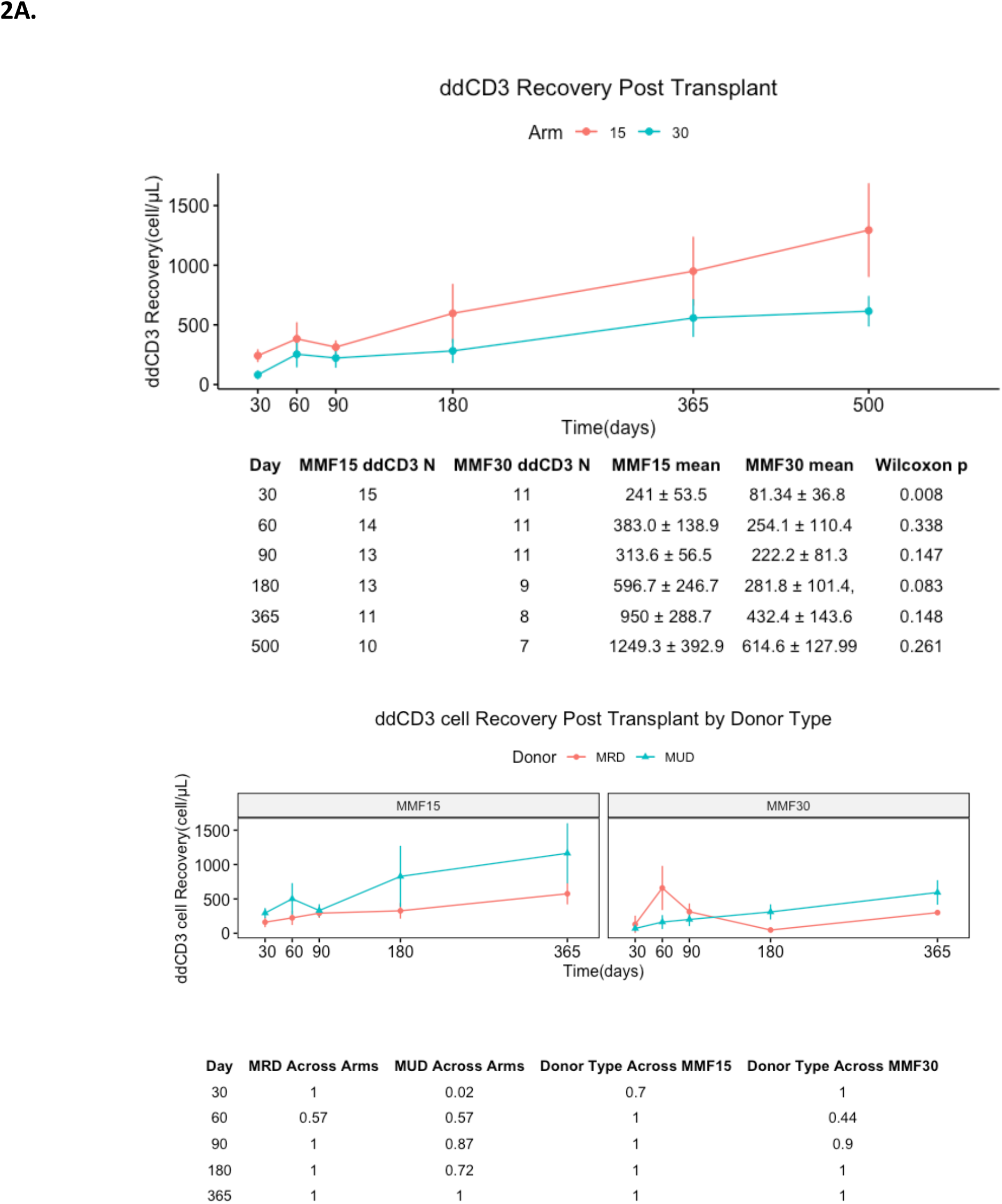
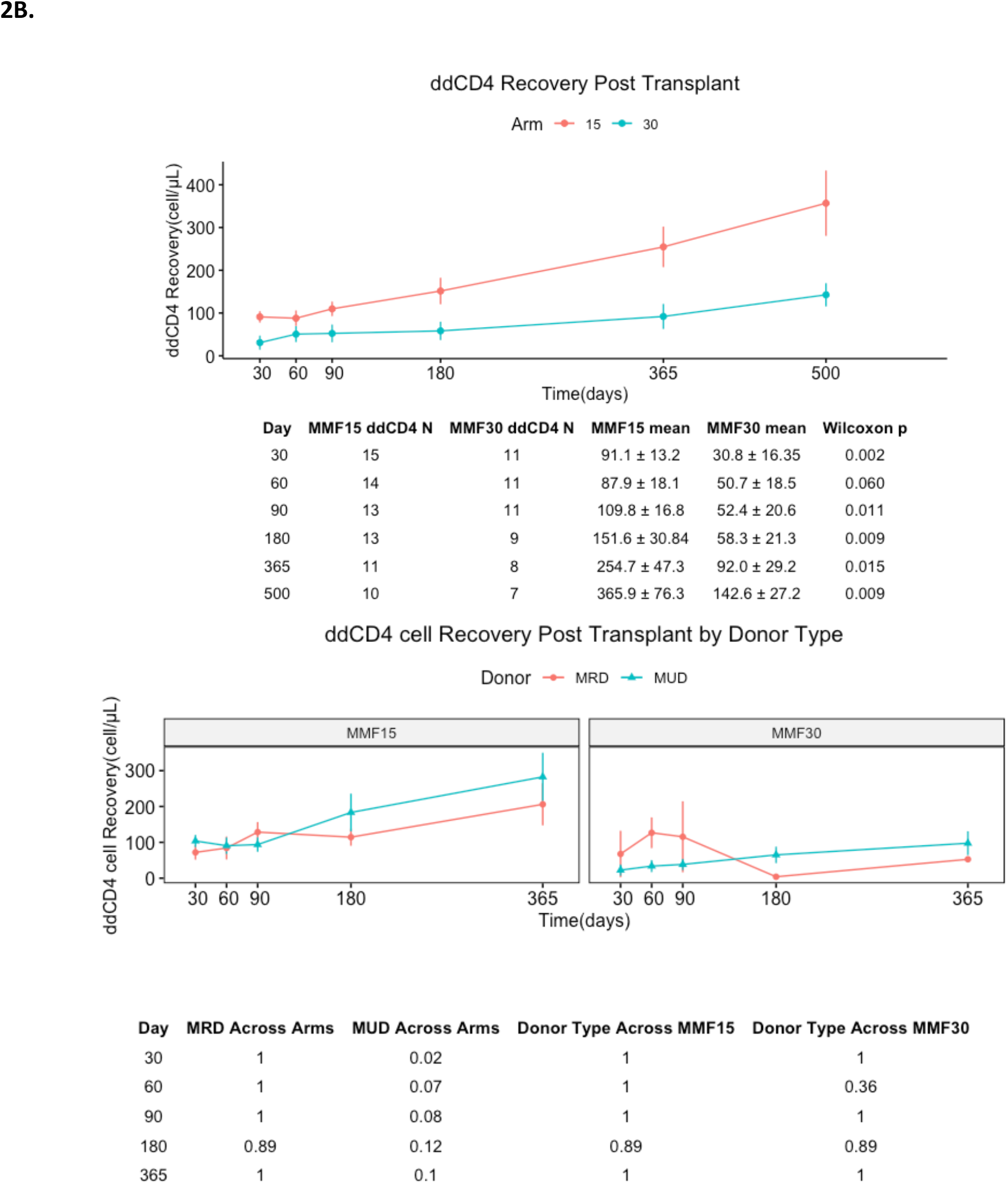
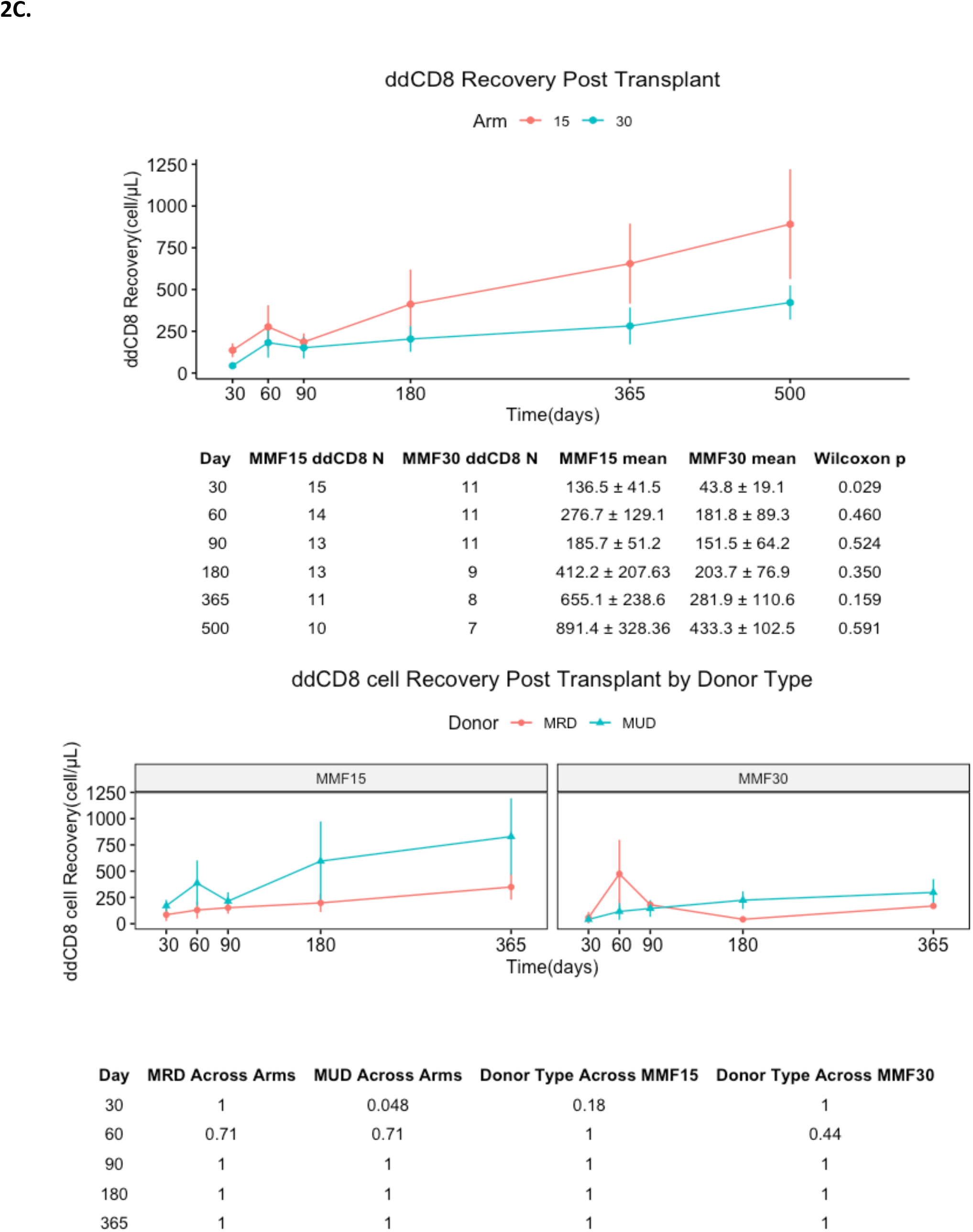
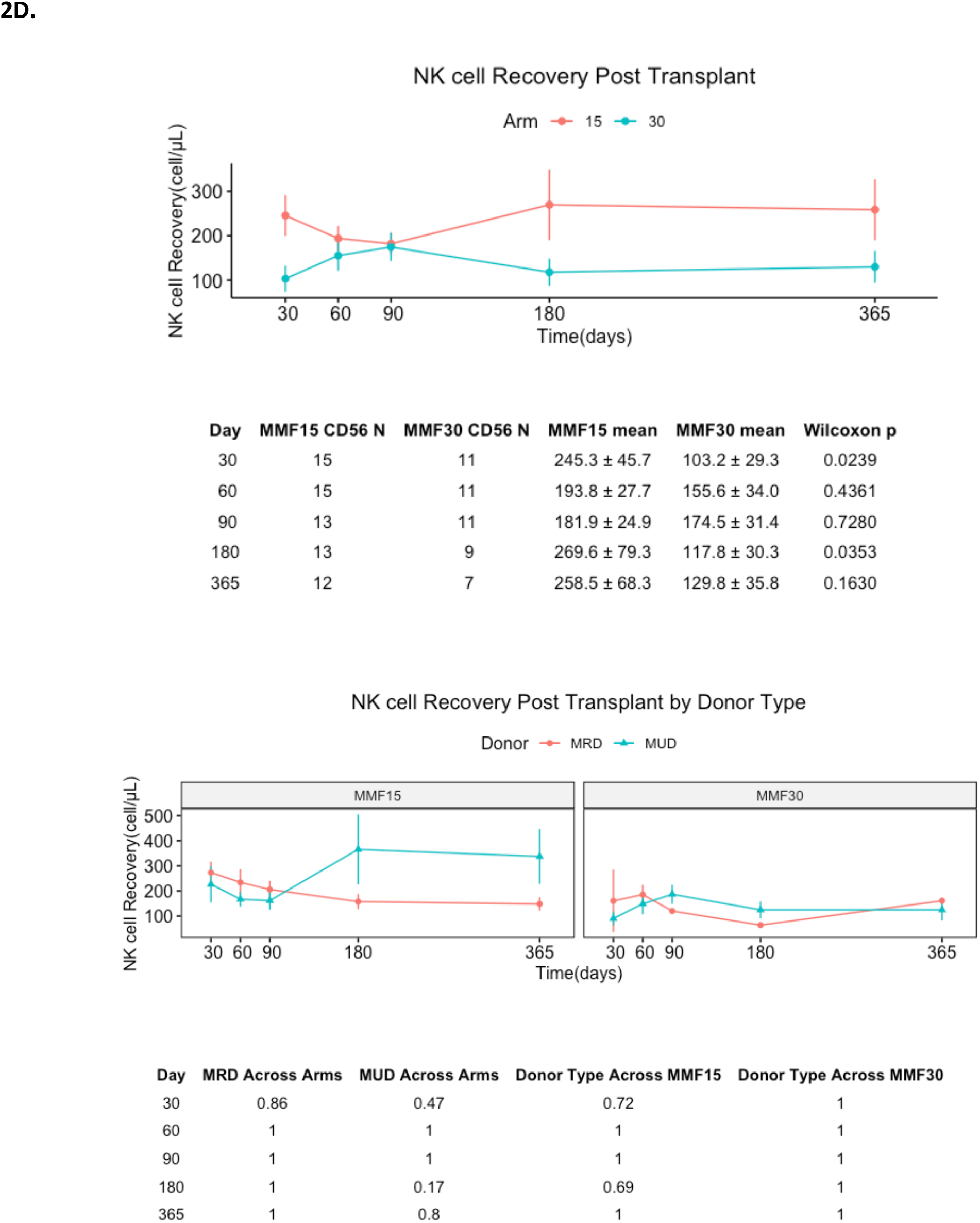
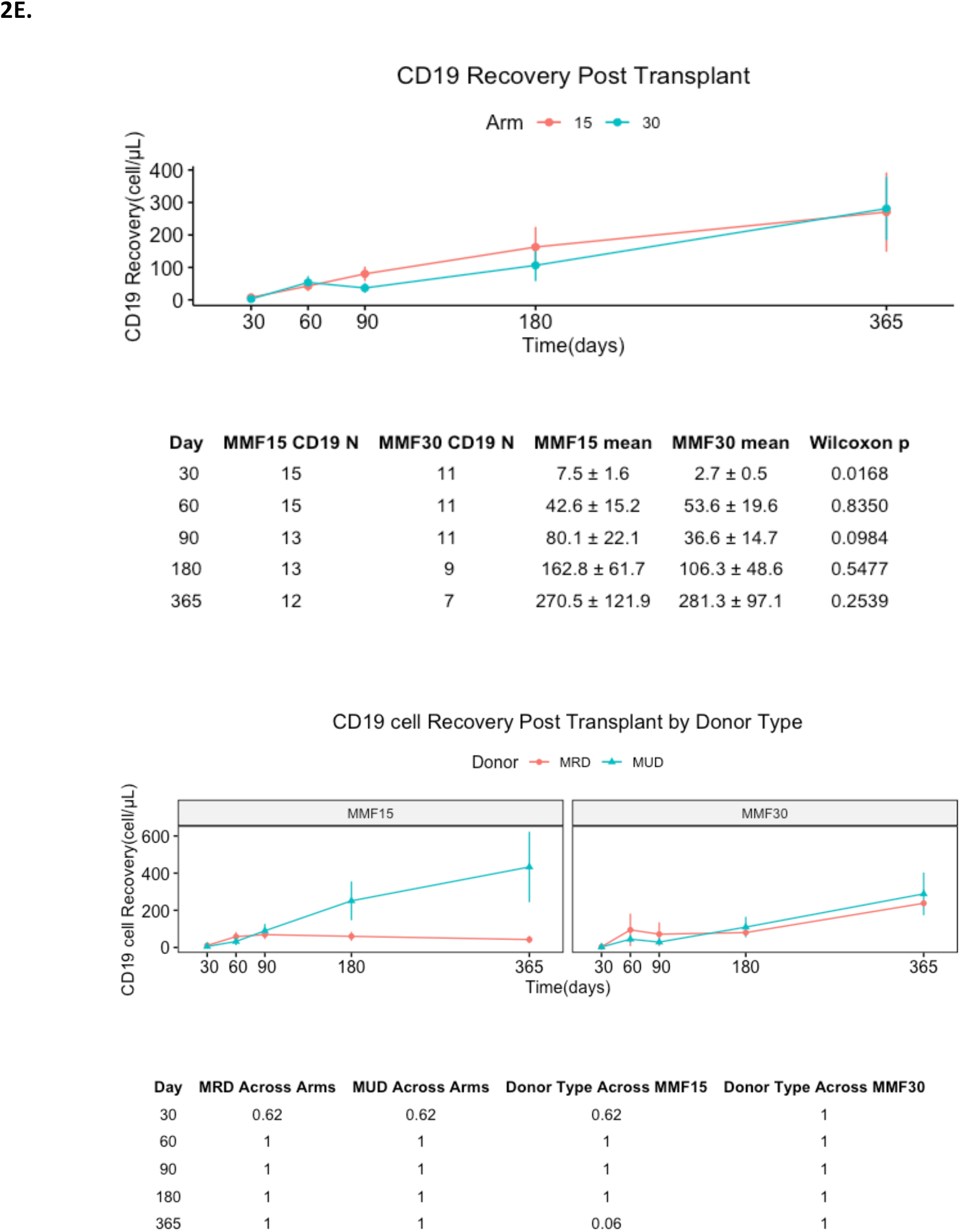
Immune cell recovery as a function of time post-transplant in MMF15 & MMF30 cohorts, and in MUD and MRD recipients. Table at the bottom of donor graphs are P values of paired Wilcoxon tests.

### Impact of immune cell recovery on clinical outcomes

The effect of T, B, and NK cell reconstitution on clinical outcomes in all trial participants was studied in the combined study population (**Supplementary Table 1**). Higher ddCD3 count on day 30 was associated with an increased risk of grade 2-4 aGVHD (HR= 1.005, p= 0.015). Of the T cell subsets, ddCD8 count on day 30 was similarly associated with increased risk of grade 2-4 aGVHD, as well as grade 3-4 aGVHD (aGVHD grade 2-4 HR; 1.008, p=0.0007, grade 3-4 HR; 1.008, p=0.04). Conversely, higher ddCD4 cell count on day 90 was associated with an increased risk of moderate to severe cGVHD (HR; 1.018, p=0.035) which corresponds to the observation of patients in the MMF15 arm having both higher CD4 counts and greater tendency to develop cGVHD (**Supplementary Figure 2**). Superior NK cell recovery on days 30 and 60 was associated with improved OS (day 30 HR; 0.9906, p;0.031, day 60 HR; 0.9886, p=0.016), and on day 90 with reduced DLI-free, relapse-free survival (HR; 1.0079, p=0.033) paralleling the trend toward improved survival and early superior NK recovery in the MMF15 arm.

### TRB repertoire evolution

TRB sequencing demonstrated that patients in the MMF15 arms (N=10 at day 30; 9 at day 100) compared with MMF30 (N=9 at day 30; 10 at day 100) consistently had a greater number of both unique, productive TRB gene rearrangements (median 26686 vs. 12892 (day 30) & 31324 vs. 15345 (day 100), P=0.11 & 0.095 respectively) and productive templates (56785 vs. 22528 (day 30) & 65458 vs. 32082 (day 100), P=NS) (**Figure 3A and 3B**). This was consistent with the observed superior numeric CD3+ T cell recovery recorded at these time points in the MMF15 cohort. Further, Simpson’s clonality calculated for productive TRB sequences was also lower for the MMF15 arm at day 100 (0.07 vs. 0.06 (day 30) & 0.07 vs. 0.12 (day 100), P=NS) indicating the evolution of a more diverse T cell repertoire in this study arm (**Figure 3C**).

**Figure 3.**
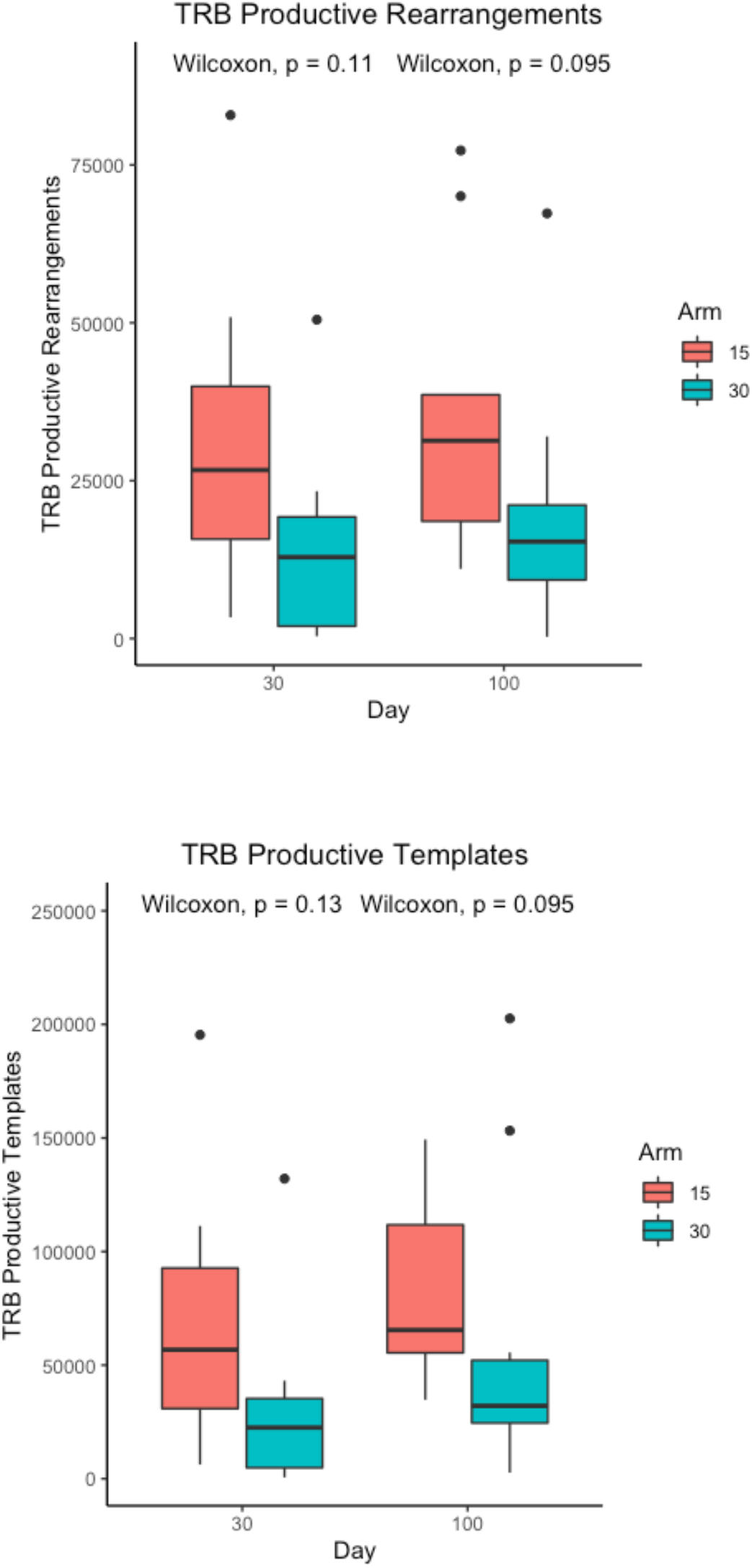
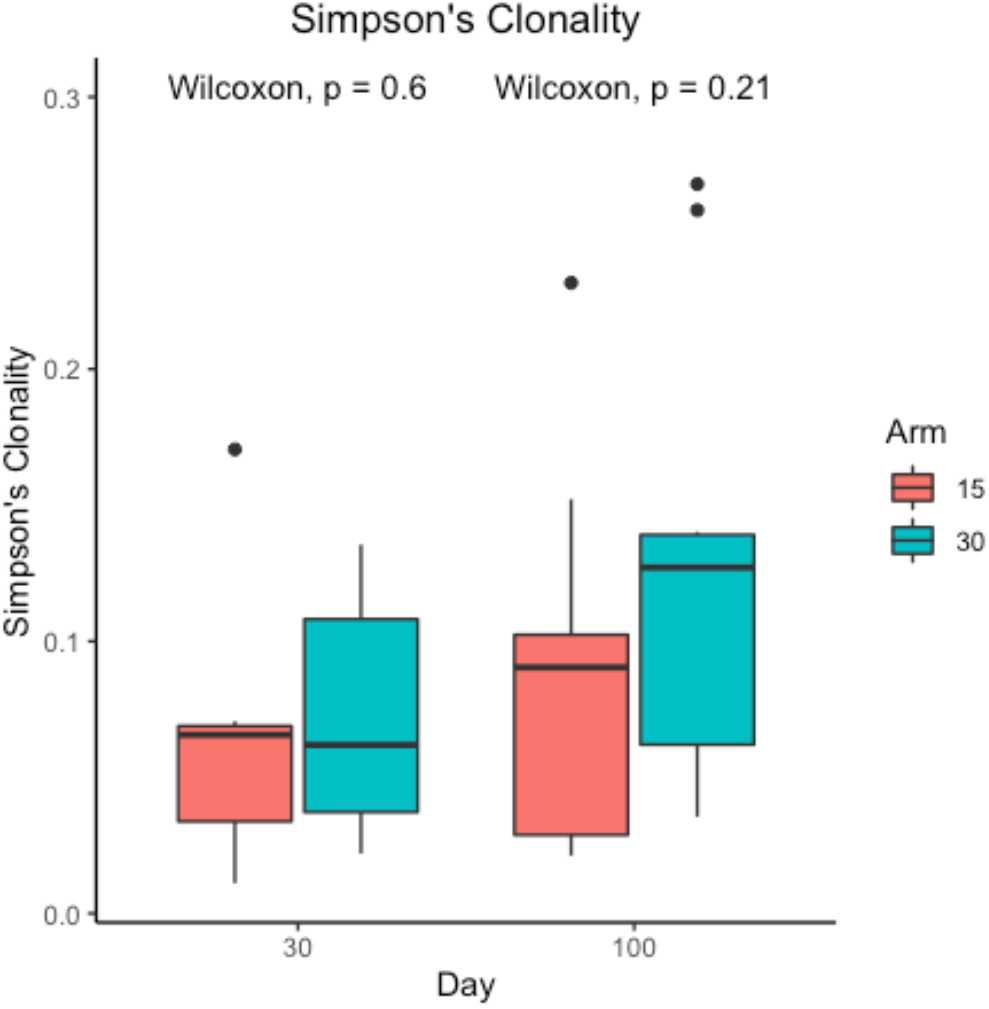
T cell receptor beta repertoire next generation sequencing. Plots depicting TRB rearrangements, TRB templates and Simpson’s clonality for MMF15 & MMF30 patients.

Productive TRB gene rearrangements and templates were plotted against simultaneously measured absolute CD3+ T cell counts in circulation, adjusted for volume of blood sequenced, to give a vector representation of the T cell repertoire at each time point for each patient. On average patients in the MMF15 arm had a vector distribution farther from the origin in the complex number plane modelling the T cell repertoire vector space (**Figure 4A**). This signified a larger and more clonally diverse T cell population. When the change in this vector from day 30 to day 100 was mapped across this plane, due to a relatively larger increase in T cell counts as opposed to number of rearrangements, these vectors transformed largely along the X-axis in most patients (**Figure 4B**). This indicated that the T cell clonal emergence is most active during the first few weeks after HCT, with growth of individual clones subsequently occurring over time. This supports the primacy of early-term immune recovery during the transplant process with defining clonal diversity emerging during that period. In this small cohort of patients, no consistent association was observed between GVHD and relapse, however probability distribution of survival may be different in the patients with different vector patterns(P=0.56) (**Figure 5**).

**Figure 4.**
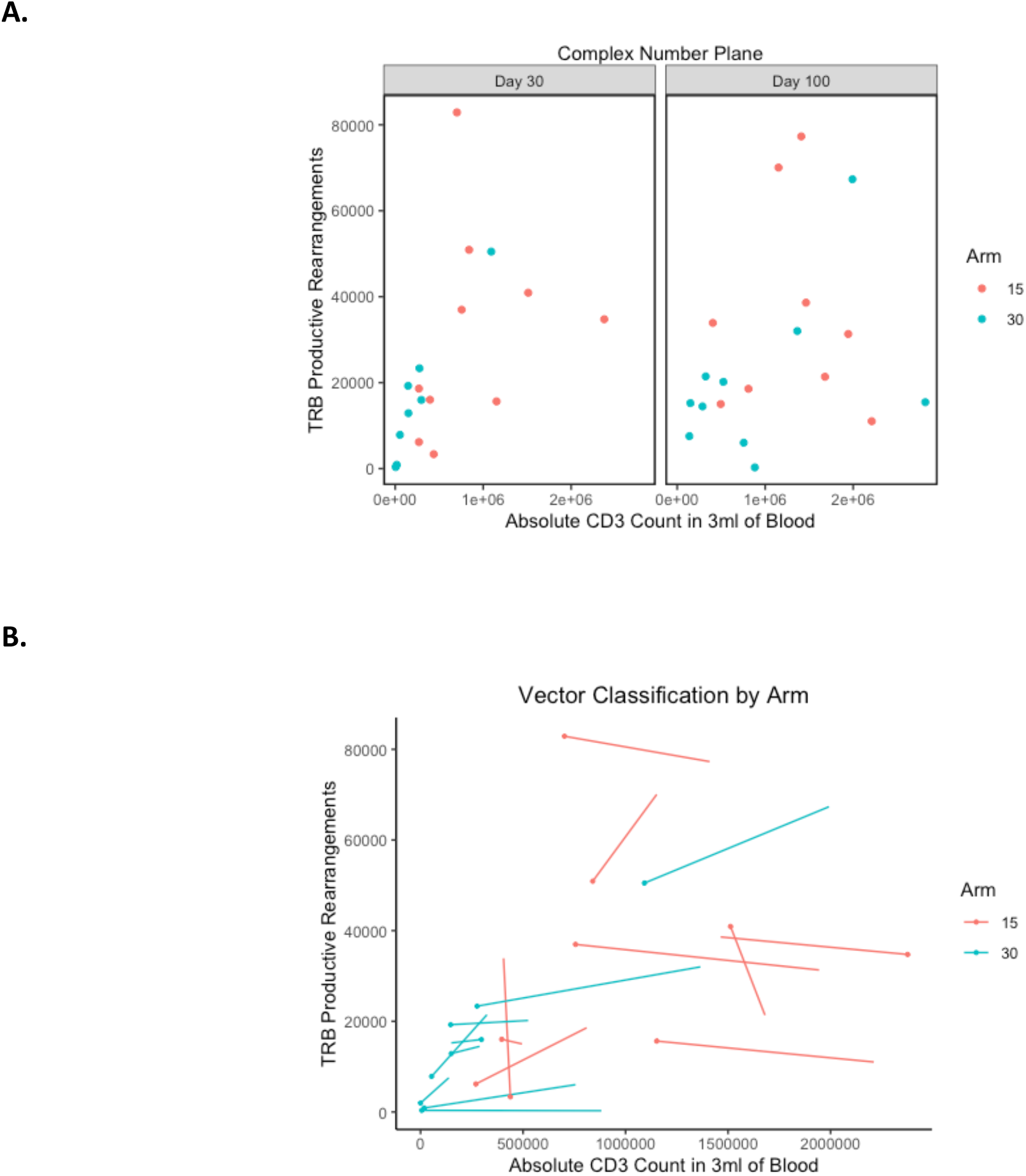
**A**. T cell repertoire vectors plotted in complex number plane, showing T cell number (x-axis) and corresponding TRB rearrangements (y-axis) at days 30 and 100 from the MMF15 (red) and 30 cohorts (blue). **B**. T cell repertoire vector transforming in individual patients over time from day 30 to day 100 (N=9 paired observations for each cohort. MMF15, Red; MMF30, Blue).

**Figure 5.**
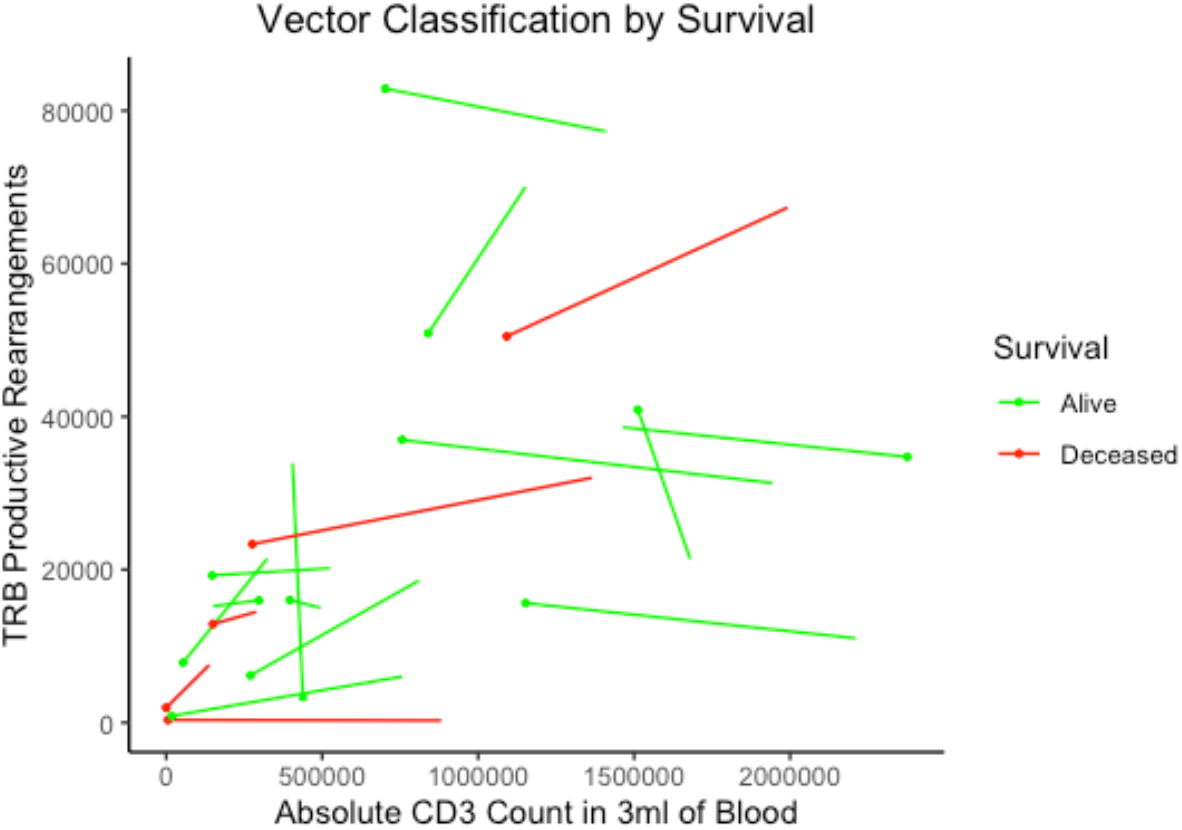
Different probability distribution of survival in different regions of the complex plane, probability of survival 0.86 vs. 0.64 (P=0.56 FET) in the area beyond the region bounded by complex number, 10^6^ + 30000 *i*, with the real (T cell count) and imaginary (TCR rearrangements) components of the T cell repertoire vector, depicted on the X and Y axes respectively.

## Discussion

In this study, recipients of allogeneic HCT underwent a uniform immunoablative conditioning regimen with rabbit ATG and reduced intensity TBI. The duration of intense post-transplant immune suppression was varied in the two study arms, as were the cytokines given for promoting engraftment. Patients who received a shorter course of intense immune suppression and GM-CSF had more rapid and sustained T cell recovery, particularly of helper T cells which continued to be higher over one year out from transplant. The T cell repertoire emerging post-transplant tended to be more diverse in this group as well. These differences in donor-derived immune recovery were associated with clinical outcome differences in the two study arms, with the expected functional consequences of greater alloreactive complications, and potential for improvement in survival in patients with superior T cell and NK cell recovery. These findings support the notion that small early differences in immune suppression, specifically, reduction in intense immune suppression duration by 15 days, led to long-term differences in immune reconstitution and had a potential for clinical outcome benefit in allograft recipients. This supports the notion that post-transplant immune recovery, with its functional implications, has dynamical underpinnings, and that, clinical outcomes following HCT are not entirely probabilistic. Adjustments to immunosuppressive therapy may be made with resulting differences in clinical outcomes, particularly in recipients of RIC regimens.

Before expanding on the findings from this study it is important to outline the shortcomings of this report. The trial was originally designed to accrue 79 patients to investigate improvement in 1-year DLI-free, relapse-free survival in the study arm. The study accrual goal could not be met because of declining trial referral. This was likely related to contemporary advances in the pharmacotherapy of relapsed lymphomas and myeloma, as well as the availability of chimeric antigen receptor T cells which reduced the desirability of allografting in these conditions. Improvements in supportive care protocols with myeloablative conditioning (MAC) regimens further reduced study referrals for myeloid disorders using this immunoablative approach. As a consequence, aside from the usual single center transplant trial issues around heterogeneity of donor and disease states, this trial is burdened with low numbers. Thus, the conclusions from this study must be interpreted keeping in mind its limitations. Nevertheless, because of adaptive randomization, the specific nature of the surrogate end point chosen (immune reconstitution at 8 weeks) and the immune reconstitution data collected, the authors posit that the T cell recovery data are robust and likely extrapolatable to future similar endeavors.

The results reported here highlight two aspects of transplant immunobiology. First, its dynamical and evolving nature as a function of time, and second, the crucial impact of the very first few days following transplant. Statistical principles and need for interpretability generally limit clinical trial designs to the study of single interventions and their effect on a desired set of clinical outcomes over the duration of the trials. This is adequate in most circumstances in oncology where a single outcome is impacted by the intervention, such as a trial of treatment response of chronic myelogenous leukemia to a tyrosine kinase inhibitor. In stem cell transplantation each intervention has a series of cascading downstream effects on the probability of multiple clinical outcomes, often of an opposite nature given the immunobiology of HCT. For example, intensifying conditioning reduces relapse, but increases TRM; conversely T cell depletion reduces GVHD, but increases relapse risk. These impacts are not instantaneous and unfold over time following HCT with varying probability. Further they are complex, with many variables influencing the physiology of immune response, including intensity and duration of immune suppression, donor-recipient whole exome variation, infections and the microbiome.

The findings reported here demonstrate the principle that immune recovery may be *modulated* with interventions in the immediate aftermath of HCT. A reduction in the duration of intense immune suppression in this instance increases the rate of T cell recovery. Risk of GVHD is increased with different T cell subsets recovering and may impact survival. As the change in therapy reported here elicits a response, ongoing adaptation to the evolving T cell milieu with therapeutic intervention may similarly influence events in the longer term. As an example, patients with an elevated CD8+ T cell count early on may benefit from an intensification of the calcineurin inhibitor therapy or introduction of additional GVHD prophylactic measures such as extra-corporeal photopheresis, anti-interleukin 6 antibodies or CD28 blockade. Patients with an elevated CD4 count may need to continue their calcineurin inhibitor for a longer period than the conventional 6 months post-transplant. The larger point to be made here is that ongoing surveillance of immune recovery and dynamic adjustment of immune suppression on an ongoing basis for many months after transplantation is imperative for reducing the risk of toxicities following HCT.

The second takeaway from these trial results is the influence that early intervention has on longer term outcomes. In this instance the MMF given over the first 15 days after HCT yielded comparable control of GVHD as the longer regimen, with improved immune recovery after discontinuing this agent. Time and again the critical impact of very early interventions following HCT, such as administration of post-transplant cyclophosphamide in haploidentical transplants, have demonstrated an enduring impact which lasts long into the course of transplant.^30^ The shorter course of MMF was chosen in this instance to mimic the time course of post-transplant methotrexate, which is administered for a shorter period, and has a significant influence on the longer-term course. In this study, early discontinuation of MMF following engraftment allowed optimal NK and T cell recovery, without an increase in risk of severe GVHD and reliable establishment of engraftment. GM-CSF was utilized in this study to augment monocyte differentiation into antigen presenting dendritic cells, and while numerically there was no difference in the magnitude of monocyte recovery between the two arms, it is expected that this cytokine would have contributed to the phenotypic maturation of antigen presenting cells^31^ and to potentially greater T cell subset numeric recovery and clonal diversity observed.^32 33^

Over the last decade TRB repertoire reconstitution as measured by next generation sequencing has emerged as a robust measure of T cell recovery. These large data sets are reported using a variety of diversity indices, such as Simpson’s clonality index and Shannon’s entropy. Other measures examining the TRB repertoire from the frame of reference of the VDJ gene segment usage, looking at relative clonal frequencies of unique clones have demonstrated repertoire properties such as fractal organization and translational symmetry. ^10 11^ Fractal structures represent iterative expansions of complex numbers and are frequently observed in nature. T cells have two unique aspects that render them effective, their ability to identify antigens through unique T cell receptors and their number. These two properties of bulk reconstituting T cell populations post-transplant, when modelled as vectors in the complex plane, demonstrate that patients in the MMF15 arm had a more numerous and diverse T cell repertoire as compared with the MMF30 arm. Importantly, it demonstrated that the period of development of greatest T cell clonal richness is the first month following transplant. Subsequent to that, the dominant effect is numeric expansion of T cells, with relatively minor change in the overall number of T cell clones. The approach of using complex numbers to study developing (fractal) T cell repertoire has value beyond simple clinical utility of predicting outcomes in a large enough cohort of patients. It provides a means for understanding *and* simulating T cell recovery in mathematical models examining immune reconstitution in different scenarios of HLA compatibility and immune suppression post-transplant. Particularly, the use of complex numbers to simulate the growing T cell repertoire in a logistic growth scenario not only accounts for the increase in T cell numbers over time, but also for the new T cell clones emerging as time passes post-transplant. This complex number representation comprehensively demonstrates the differences in TRB repertoire emerging in the two study arms (**Figure 4B**). Indeed, it may be utilized to study the differences in immune cell repertoire recovery between patients getting different immune suppressive therapies. The complex plane provides a new method of tracking immune reconstitution and probability density distribution of clinical events post-transplant in a large enough cohort of patients (**Figure 5**).

NMA and RIC HCT is complicated by high rates of GVHD.^34^ T cell depletion with ATG reduces the risk of chronic GVHD in RIC HCT,^35^ however if infections, autologous reconstitution and recurrent disease are to be minimized,^36^ optimizing post-engraftment immune suppression is critical. The original MMF-containing regimens were critical in preventing both GVHD and graft rejection, and while the former is mitigated with ATG, the autologous T cell recovery may be seen late after transplant when ATG is used. Reducing intense immune suppression in the early days following transplantation would counter this by improving early donor T cell recovery. Reduction in the duration of myelosuppression from MMF likely has a synergistic effect of the innate-adaptive immune interaction, particularly in recipients of GM-CSF. These effects will require more detailed immunophenotypic characterization, because in this study measuring simple blood counts, there was a significant difference in the engraftment kinetics of patients getting GCSF vs. GM-CSF. Faster myeloid recovery in the former was likely related to the cytokine utilized, and lymphoid recovery in the latter, related to duration of MMF therapy as well as the dendritic cell differentiation of monocytes.

In conclusion, the risk of compromised immune recovery in recipients of ATG in RIC HCT may be overcome by reducing the duration of intense immune suppression post-transplant and by utilizing GM-CSF. Patients with immune recovery have optimal outcomes, evident even in a small, heterogenous cohort of HCT recipients. Constant measurement of immune reconstitution and adjustment of immune suppression following HCT is a necessary condition for optimizing clinical outcomes.

## Data Availability

All data produced in the present work are contained in the manuscript

## Acknowledgements

The authors gratefully acknowledge bone marrow transplant coordinators and nurses in the Cellular Immunotherapy and Transplant Program at VCU. The Study was supported by Massey Cancer Center; AT was supported by research funding from the NIH-NCI Cancer Center Support Grant (P30-CA016059; PI: Gordon Ginder, MD).

## Author contributions

VZ, Data collection and statistical analysis, wrote the paper; MS, TRB sequencing and analysis; GB, TRB sequencing and analysis; SW, Data collection and analysis; AY, Data collection and analysis; TA, Data collection and analysis; GS, critical review and edit manuscript; EK, critical review and edit manuscript; MA, critical review and edit manuscript; JR, Critical review and edit manuscript; CR, patient enrollment, study coordination, data management; RS, Study design, Bayesian adaptive randomization, statistical analysis and wrote manuscript; AT, study design, mathematical analysis, data review and analysis, wrote the manuscript.

## Supplementary Materials

**Supplementary Figure 1.**
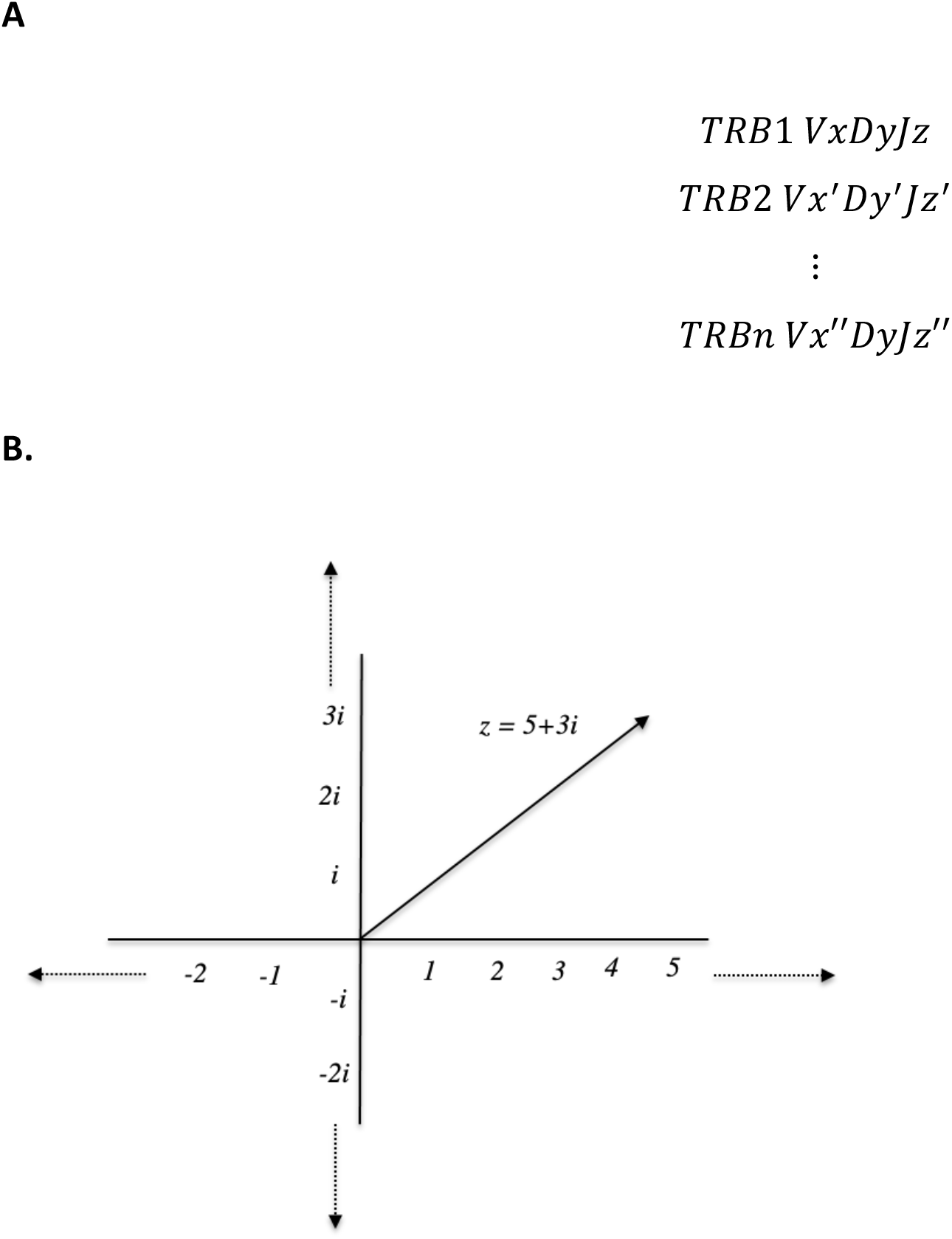
**A**. Matrix illustrating multiple T cell clones (TRB1 …. n) with unique VDJ rearrangements, constituting the TRB multicomponent vector. **B**. Complex number plane to plot complex valued vectors. Real component plotted on the x-axis, with the imaginary component plotted on the y-axis. Complex numbers are used in calculations, depicting real number value (e.g., *5*), such as CD3+ T cell counts, with the imaginary component (e.g., *3i*) in this instance representing a quality metric, such as clonality for the population of T cells depicted. In this example, the T cell repertoire vector is comprised of 5 T cells, made up of 3 distinct clones.

**Supplementary Figure 2.**
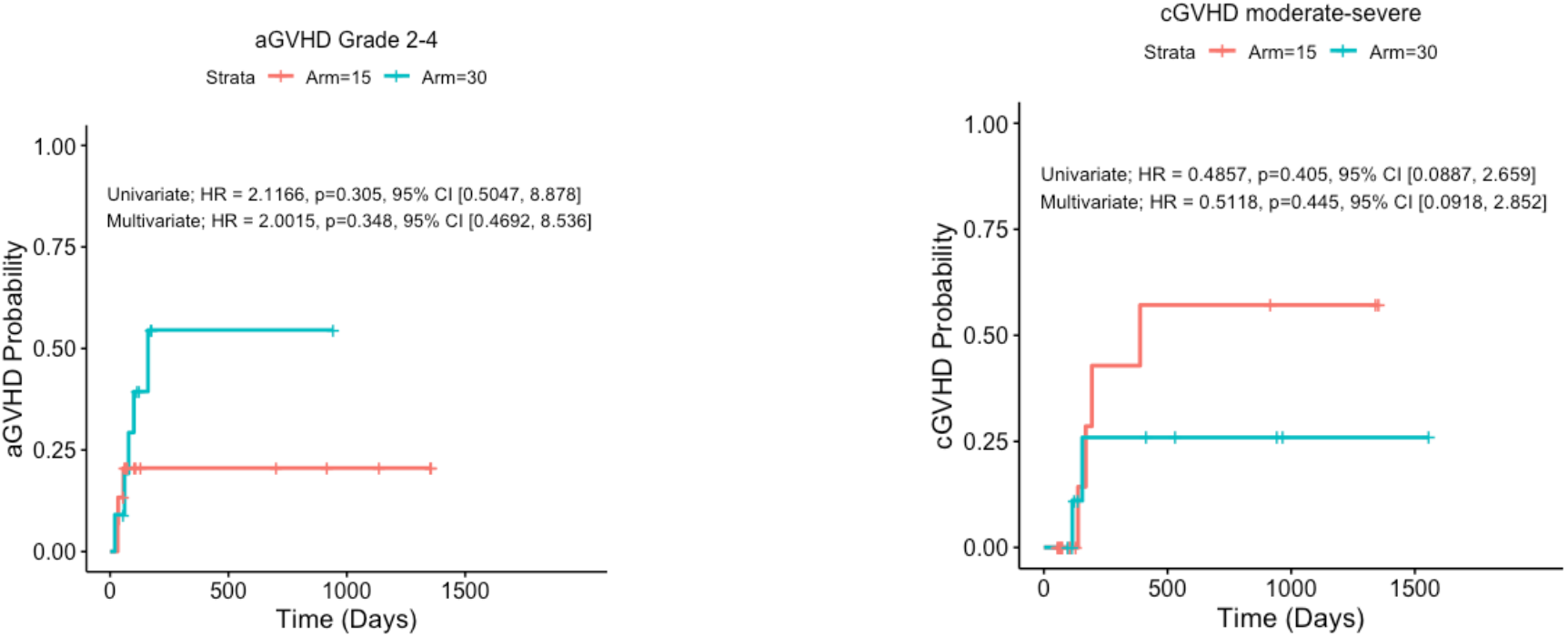
Cumulative incidence of acute and chronic GVHD.

**Supplementary Figure 3.**
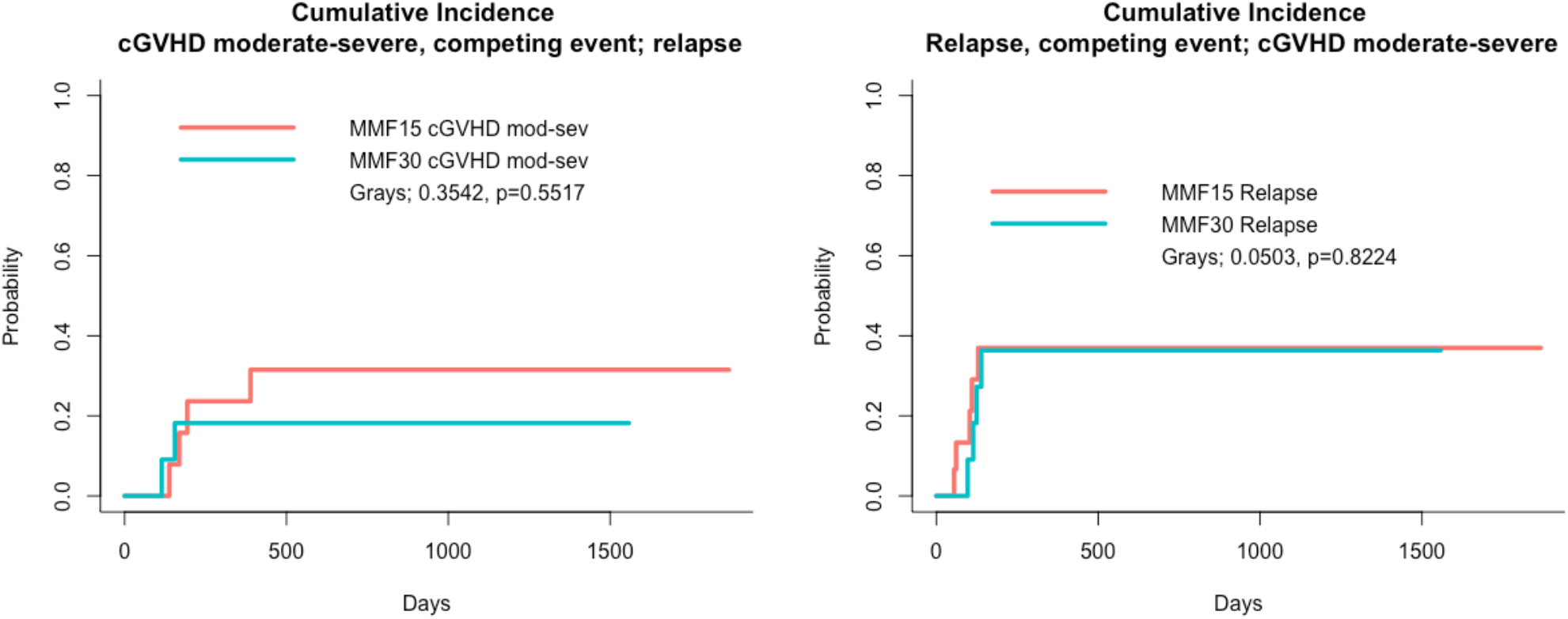
Cumulative incidence of relapse and chronic GVHD.

**Supplementary Table 1.** Multivariate analysis of the impact of immune recovery over time on clinical outcomes.

|  |  | Survival |  | Relapse |  | DLI-free Relapse-Free Survival |  | aGVHD Grade 2-4 |  | aGVHD Grade 3-4** |  | cGVHD Moderate-Severe |  | cGVHD Severe** |  |
| --- | --- | --- | --- | --- | --- | --- | --- | --- | --- | --- | --- | --- | --- | --- | --- |
| Cell Type | Day | Hazard Ratio | P value | Hazard Ratio | P value | Hazard Ratio | P value | Hazard Ratio | P value | Hazard Ratio | P value | Hazard Ratio | P value | Hazard Ratio | P value |
| ddCD4 | 30 | 0.993 | 0.371 | 0.995 | 0.483 | 0.996 | 0.434 | 1.004 | 0.572 | 0.999 | 0.890 | 1.008 | 0.338 | 1.007 | 0.515 |
|  | 60 | 0.988 | 0.140 | 0.993 | 0.264 | 0.992 | 0.173 | 1.006 | 0.283 | 0.996 | 0.579 | 1.008 | 0.260 | 1.008 | 0.398 |
|  | 90 | 1.002 | 0.731 | 1.002 | 0.699 | 1.003 | 0.526 | 1.001 | 0.854 | 0.997 | 0.735 | 1.018 | 0.035 | 1.019 | 0.120 |
| ddCD8 | 30 | 0.999 | 0.801 | 0.989 | 0.201 | 0.997 | 0.253 | 1.008 | 0.007 | 1.008 | 0.040 | 1.001 | 0.699 | 1.002 | 0.616 |
|  | 60 | 0.992 | 0.212 | 1.000 | 0.785 | 0.999 | 0.395 | 1.001 | 0.220 | 1.000 | 0.864 | 0.995 | 0.186 | 0.997 | 0.435 |
|  | 90 | 0.997 | 0.364 | 0.999 | 0.684 | 0.998 | 0.447 | 1.002 | 0.185 | 0.999 | 0.739 | 0.998 | 0.513 | 0.998 | 0.568 |
| ddCD3 | 30 | 0.999 | 0.615 | 0.995 | 0.178 | 0.998 | 0.230 | 1.005 | 0.015 | 1.004 | 0.077 | 1.001 | 0.563 | 1.002 | 0.530 |
|  | 60 | 0.995 | 0.083 | 0.999 | 0.584 | 0.999 | 0.267 | 1.001 | 0.176 | 1.000 | 0.845 | 0.998 | 0.311 | 0.999 | 0.519 |
|  | 90 | 0.999 | 0.455 | 0.999 | 0.762 | 0.999 | 0.575 | 1.001 | 0.246 | 0.999 | 0.730 | 1.000 | 0.970 | 1.000 | 0.943 |
| CD19 | 30 | 0.994 | 0.942 | 0.948 | 0.575 | 1.045 | 0.549 | 1.102 | 0.346 | 1.068 | 0.533 | 0.979 | 0.887 | 1.052 | 0.790 |
|  | 60 | 0.988 | 0.103 | 1.000 | 0.931 | 0.998 | 0.723 | 0.997 | 0.690 | 1.004 | 0.576 | 1.009 | 0.200 | 1.020 | 0.067 |
|  | 90 | 0.980 | 0.076 | 0.995 | 0.429 | 0.994 | 0.277 | 1.009 | 0.137 | 1.021 | 0.035 | 1.008 | 0.127 | 1.036 | 0.272 |
| CD56 | 30 | 0.991 | 0.031 | 1.001 | 0.621 | 1.000 | 0.865 | 1.001 | 0.682 | 1.002 | 0.577 | 0.991 | 0.131 | 0.995 | 0.497 |
|  | 60 | 0.989 | 0.016 | 0.998 | 0.660 | 0.995 | 0.179 | 0.999 | 0.699 | 1.000 | 0.981 | 1.010 | 0.196 | 1.024 | 0.170 |
|  | 90 | 1.004 | 0.386 | 1.007 | 0.093 | 1.007 | 0.039 | 0.995 | 0.224 | 0.999 | 0.878 | 1.004 | 0.529 | 1.003 | 0.734 |
| ** Model did not converge due to "donor type" variable and analysis was performed with the variables omission. |  |  |  |  |  |  |  |  |  |  |  |  |  |  |  |

